# Impact of COVID-19 Pandemic on Inpatient Rehabilitation and the Original Infection Control Measures for Rehabilitation Team

**DOI:** 10.1101/2021.01.20.21250145

**Authors:** Yukimasa Igawa, Takahiro Sugimoto, Hiromasa Horimoto, Yusuke Moriya, Masaki Okada, Yasuyuki Yamada

**Affiliations:** Department of Physical Medicine and Rehabilitation, Aizen Hospital, Sapporo, Hokkaido, Japan; Department of Rehabilitation, Aizen Hospital, Sapporo, Hokkaido, Japan; Department of Neurology, Aizen Hospital, Sapporo, Hokkaido, Japan

**Keywords:** COVID-19, Rehabilitation Hospital, Outcome Assessment, Nosocomial Infection

## Abstract

**Objective:** This study aimed to investigate the impact of the coronavirus disease 2019 (COVID-19) pandemic on inpatient rehabilitation, and to determine the effectiveness of the original infection control measures implemented for the rehabilitation team.

**Methods:** In this single-center, retrospective, observational study, we calculated multiple rehabilitation indices of patients discharged from our rehabilitation ward between February 28 and May 25, 2020 when Hokkaido was initially affected by COVID-19, and compared them with those calculated during the same period in 2019. Fisher’s exact test and the Mann-Whitney U test were used for statistical analysis. We also verified the impact of implementing the original infection control measures for the rehabilitation team on preventing nosocomial infections.

**Results:** A total of 93 patients (47 of 2020 group, 46 of 2019 group) were included. The median age was 87 and 88 years, respectively, with no differences in age, sex, and main disease between the groups. Training time per day in the ward in 2020 was significantly lower than that in 2019 (p = 0.013). No significant differences were found in the qualitative evaluation indices of Functional Independence Measure (FIM) score at admission, FIM gain, length of ward stay, FIM efficiency, and rate of discharge to home. None of the patients or staff members had confirmed COVID-19 during the study period.

**Conclusions:** Early COVID-19 pandemic in Hokkaido affected the quantitative index for inpatient rehabilitation but not the qualitative indices. No symptomatic nosocomial COVID-19 infections were observed with our infection control measures.

## Introduction

In December 2019, an emerging infectious disease caused by severe acute respiratory syndrome coronavirus 2 (SARS-CoV-2) was reported from China,^1)^ and the World Health Organization declared coronavirus disease 2019 (COVID-19) a global pandemic on March 11, 2020.^2)^ In Japan, the first case of COVID-19 was reported in January 2020.^3)^ Since then, the number of reported cases had continued to increase and Hokkaido had become the most heavily impacted area in Japan. Taking the situation very seriously, the Hokkaido Government issued a “Declaration of State of Emergency in Hokkaido” on February 28, 2020.^4)^ With a decrease in the number of reported cases, Hokkaido’s declaration of a state of emergency was then lifted on March 19. However, the number of reported infections increased again across the country, including Hokkaido area, in early April, and the prime minister of Japan issued a nationwide declaration of a state of emergency on April 16. This increase was considered to be the second wave of COVID-19 in Hokkaido, which had not been seen nationally or globally. The situation under the declaration of a state of emergency in Hokkaido continued until May 25, when it was lifted nationwide.^5)^ The early spread of the COVID-19 epidemic and the second wave of outbreaks in Hokkaido had a great impact on medical institutions and their medical staff, as well as hospitalized patients and families. Several facilities with nosocomial COVID-19 infections were reported.

Rehabilitation medicine inevitably involves close contact with patients and provides many opportunities to meet with patients and their family members to assist with hospital discharge. We hypothesized that the original infection control measures implemented for the rehabilitation team would enable both prevention of nosocomial infections and allow usual outcomes of rehabilitation medicine. To test this hypothesis, we investigated the impact of COVID-19 pandemic on inpatient rehabilitation and the effectiveness of the original infection measures implemented for the rehabilitation team.

## Materials and Methods

### Study design

Single-center, retrospective, observational study.

### Study facility

Rehabilitation ward of a hospital in the central city of Hokkaido, Japan, which did not treat patients with COVID-19.

### Subjects and evaluation items

The detailed evaluation indices obtained from discharged patients between February 28, 2020, and May 25, 2020, were compared with those obtained for the same period in 2019. The study period in 2020 started from the declaration of Hokkaido’s state of emergency and ended with the lifting of the nationwide state of emergency. Patients who were transferred to another hospital or ward because of worsening condition or adverse events and the ones that expired during the study period were excluded. We used the Functional Independence Measure (FIM) to measure changes in functional ability. The tool consists of 18 items, each of which is a 7-point ordinal scale.^6)^ The evaluation items were age, sex, main disease name, FIM score at admission, gain of FIM, length of ward stay, FIM efficiency, rate of discharge to home, and training time per day. Discharge to home was defined as discharge to the patient’s own residence or residential facility.

### Statistical analyses

The Fisher exact test was used to analyze sex, main disease, and rate of discharge to home of the subjects. The Mann-Whitney U test was used to analyze the subjects’ age, FIM score at admission, FIM gain, FIM efficiency, and training time. The level of significance was set at p < 0.05, and the statistical software program used was EZR.^7)^

### Nosocomial infection control

The infection control committee in the hospital formulated nosocomial infection control measures based on the knowledge provided by related academic societies and public institutions. The nature of rehabilitation medicine was also taken into consideration, with rehabilitation staff meetings in the hospital to consider the formulation of the original infection control measures for the rehabilitation team. Table 1 shows the infection control measures for the rehabilitation team assembled by the staff. The handling of suspected cases and polymerase chain reaction (PCR) testing for COVID-19 was supposed to be carried out by a local governmental healthcare center upon request from attending clinicians, based on the guidelines of the central government.^8)^

**Table 1:**
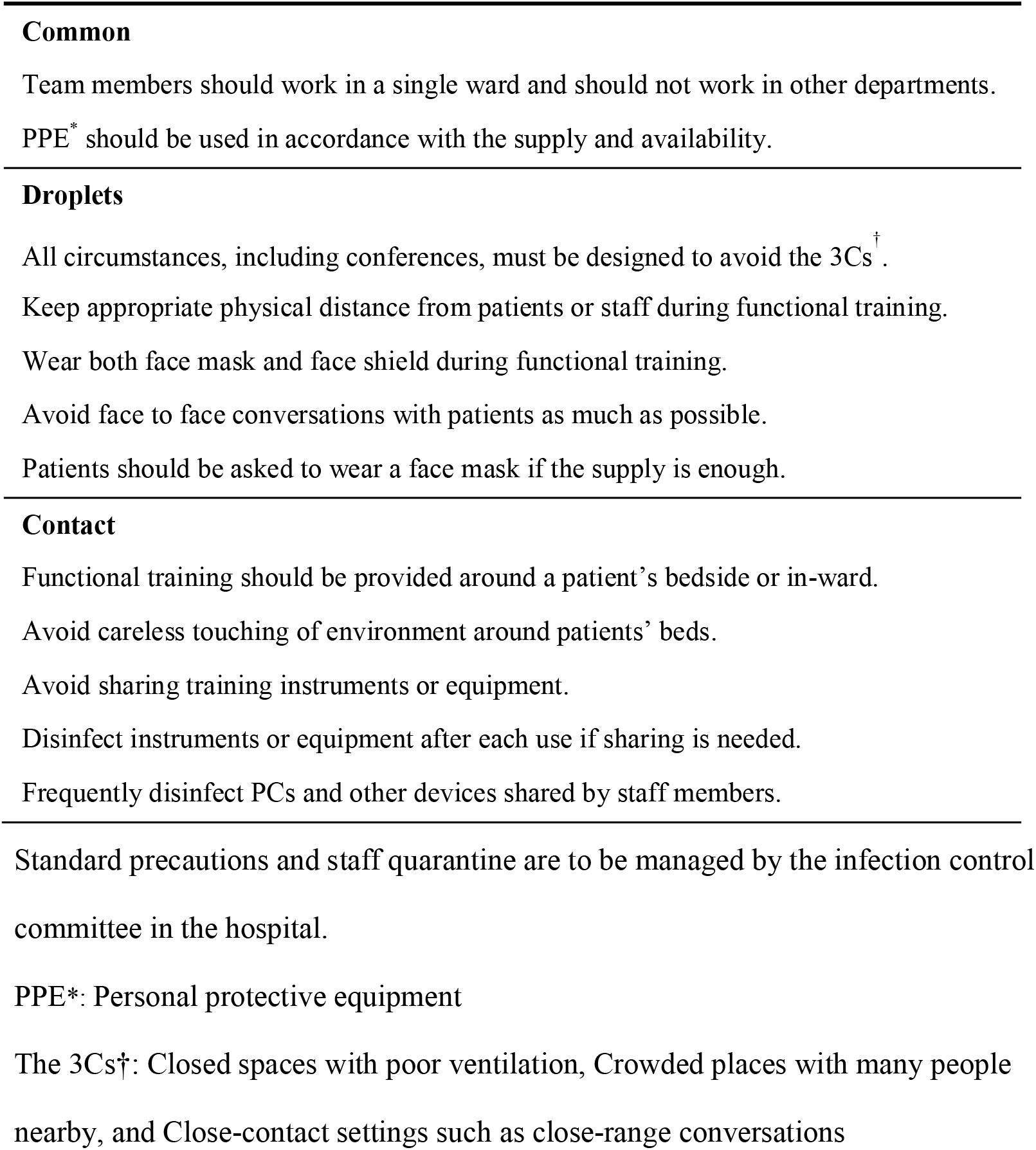
Infection control measures for the rehabilitation team.

### Ethical considerations

This was a retrospective, observational study, conducted in accordance with the tenets of the Declaration of Helsinki, guaranteeing complete anonymity. This study was approved by the Ethics Committee of Aizen Hospital (permission number: 2020-003).

## Results

### Evaluation indices measured in discharged patients

Fifty-four patients in 2020 and 58 in 2019 were discharged from the rehabilitation ward. Forty-seven study subjects in 2020 and 46 in 2019 ultimately met the inclusion criteria. Table 2 shows the demographic information for the subjects, their main diagnoses, and their evaluation index data. No significant differences in demographics and diagnoses were observed between the two groups (p > 0.05). No significant differences were found in FIM score at admission, FIM gain, length of ward stay, FIM efficiency, or rate of discharge to home (p > 0.05). Only the training time per day showed a significant decrease (p = 0.013) in the 2020 group compared with that in the 2019 group. Figure 1 shows a dot plot of the distribution of the training time per patient per day. The distribution of dots in the 2020 group was moved down compared with that in the 2019 group.

**Table 2:** Basic information and evaluation indices for discharged patients.

|  | 2020 | 2019 | *p*<sup>¶</sup> |
| --- | --- | --- | --- |
| n | 47 | 46 |  |
| Age [years]<sup>‡</sup> | 87 (80.5-91) | 88 (82.3-92.8) | 0.249 |
| Male female | 13 34 | 17 29 | 0.38 |
| CNS/Orthopedics/other<sup>§</sup> | 11/35/1 | 13/33/0 | 0.726 |
| \|\|FIM at admission<sup>‡</sup> | 61 (49.5-80.5) | 64 (46.5-79.5) | 0.982 |
| \|\|FIM gain<sup>‡</sup> | 26 (17.5-35 ) | 26 (10.3-34.8) | 0.569 |
| Length of ward stay<sup>‡</sup> | 63 (47-96.5) | 69.5 (49.3-82.8) | 0.833 |
| \|\|FIM efficiency<sup>‡</sup> | 0.47 (0.2-0.64) | 0.44 (0.19-0.62) | 0.429 |
| Rate of discharge to home | 80.9% | 89.1% | 0.762 |
| Training time per day [min]<sup>‡</sup> | 144.6 (122.3-155.4) | 155.7 (138.9-165.4) | 0.013 |

‡Numbers are given as median (interquartile range).
§Central nervous system: cerebrovascular disease or spinal cord disease. Orthopedics: bone fracture or ligament injury or disc herniation. Other: disuse syndrome.
||FIM: Functional independence measure
¶The P values for sex, main disease, and rate of discharge to home were calculated using the Fisher exact test. Age, FIM at admission, FIM gain, FIM efficiency, and training time were calculated using the Mann-Whitney U test.

**Figure 1.**
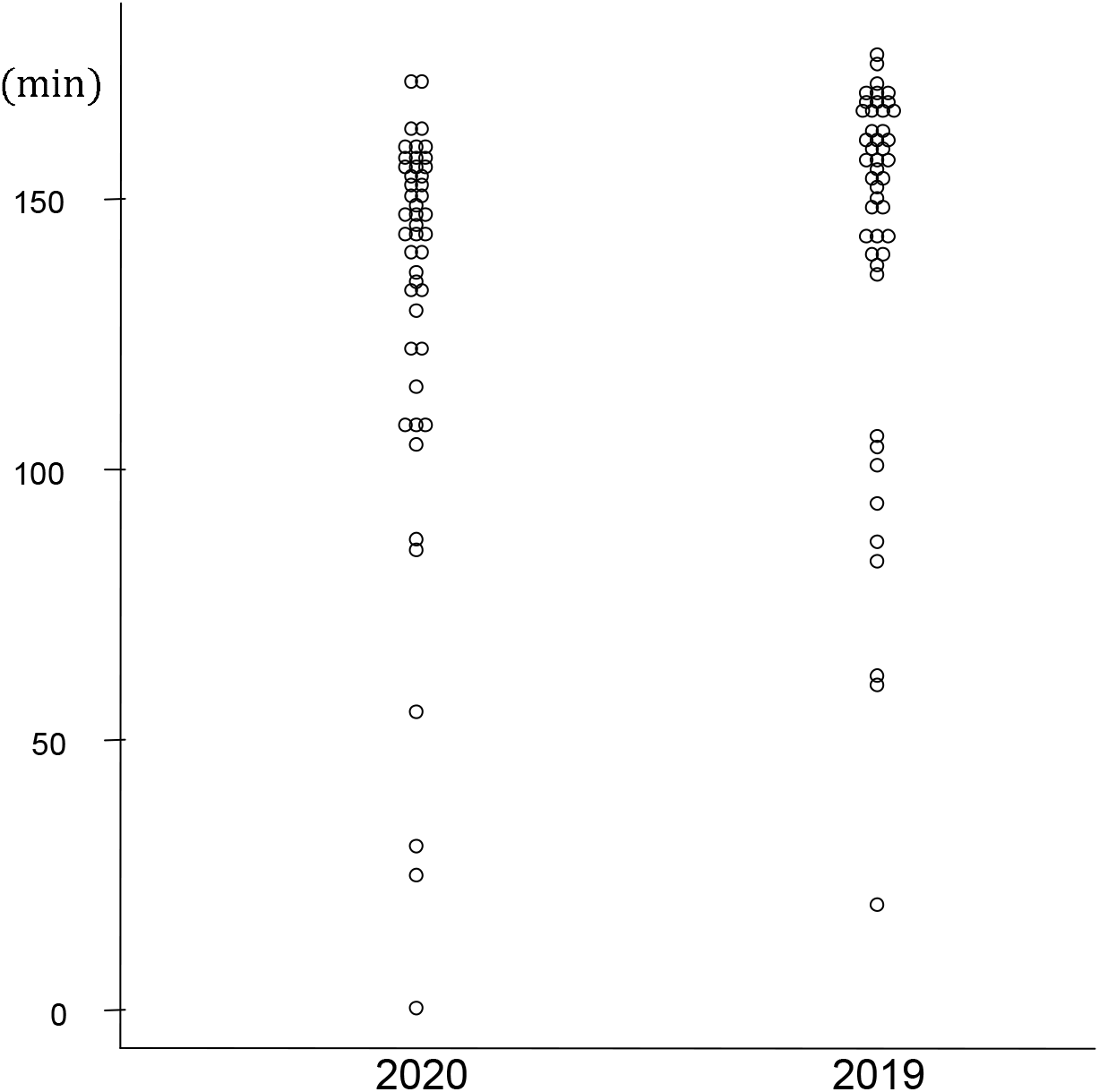
Training time of discharged patients. The distribution of training time of a discharged patient per day is shown.

### Nosocomial infections

No patients or staff members in the hospital had confirmed or suspected cases of COVID-19 during the study period. None of the staff members were eligible for PCR test based on case definitions in the guidelines, including the criteria for consultation and reporting to the local governmental healthcare center.

## Discussion

Rehabilitation outcomes have often been measured with evaluation indices based on FIM, which were designed to quantify the degree of disability.^6,9)^ Adding length of ward stay, rate of discharge to home, and training time as the evaluation indices in this study allowed a more detailed and multi-layered assessment. FIM gain is calculated by subtracting the FIM score at admission from the FIM score at discharge, and FIM efficiency is calculated by dividing the FIM gain by the number of days in the ward, all of which are indicators of the quality of inpatient rehabilitation.^10)^

If the pandemic had an impact on the quality of inpatient care, some changes in FIM-based indices and hospital length of stay should have been observed. However, the 2020 group had the same length of ward stay, unchanged FIM gain, and increased FIM efficiency. The indices obtained from the discharged patients during the study period showed no significant differences, except for the training time, between the 2020 and 2019 groups. The possible reasons for the statistically significant decrease in training time include a thorough staff quarantine under the state of emergency due to the COVID-19 outbreak, where rehabilitation therapists with the slightest change in health conditions such as the emergence of cold symptoms were instructed to stay home. Most countries, including Japan, provide guidelines for identifying healthcare workers with suspected COVID-19, which can restrict their attendance based on self-assessments including fever.^11)^ These staffing limitations might have affected the capability to provide training time during the study period. The sudden suspension of rehabilitation therapists from work appears to have had the effect of pushing down the distribution of dots per patient per day in 2020 (Fig.1). However, it is not possible to determine from this study whether the statistically significant reduction in training time had any clinical significance. Even so, the fact that COVID-19 pandemic had a significant impact on training time in inpatient rehabilitation indicates that in the uncertain future of this pandemic, telemedicine and self-training must be enhanced to complement the reduced training time.^12)^

None of the patients or staff members were observed with clinically apparent nosocomial COVID-19 during the study period. Nosocomial infection control is a top priority during the COVID-19 outbreak. Centers for Disease Control and Prevention published successive updates to guidelines intended for medical institutions.^13)^ However, there was little mention of rehabilitation medicine or facilities in the guidelines. This had led the physicians and administrators involved to develop their own infection control measures. Infection control in rehabilitation medicine is complicated by the considerable amount of contact with patients, close physical proximity, frequency of conversations, frequent production of droplets during speech therapy training or eating and swallowing training, frequent crowding among staff at conferences, and sharing of instruments and equipment. Estimates of the route of infection with COVID-19 have reportedly identified that approximately 10% of infections are contracted through environments such as high-frequency contact surfaces.^14)^ In addition to the recommended infection control measures for COVID-19 from most of the public institutes, additional measures addressing contact with equipment are required in rehabilitation medicine, where the frequency of such contact is high.

The use of surgical masks and face shields by healthcare professionals has previously been recommended and reviewed, mainly with the purpose of protecting healthcare professionals’ own body from exposure to sources of infection.^15)^ In the case of COVID-19, however, non-first-line healthcare workers had a higher infection rate than first-line healthcare workers, which differed from the observations in the previous viral disease epidemics.^16)^ In the clinical setting of rehabilitation medicine, young medical staff providing care to older patients is quite common. The older the patient and the more chronic diseases they have, the more vulnerable they are to the virus, ^17)^ and therefore, protecting hospitalized older adults from staff-mediated infections has been an urgent issue.^18)^ The concept of source control was proposed to address this issue, and global advocation for universal face masking had emerged.^19)^ For face shields, verification of the assumed influenza infection cases that were reported revealed a 96% decrease in the risk of droplet inhalation.^20)^ Although the effectiveness of using face shields in COVID-19 remains unclear, it is common sense to wear them for protection. Even in the context of an extreme shortage of Personal Protective Equipment (PPE), face shields can be handmade from commonplace materials, making them an option for universal use, as well as for use by non-first-line medical care workers,^21)^ including those in rehabilitation medicine.

The infection control measures implemented based on these findings have allowed us to continue our inpatient care without a single case of infection in an inpatient or staff member from before the declaration of the state of emergency to November 2020.

In the epicenter of the global pandemic, such as the United States and Europe, some rehabilitation wards were converted to COVID-19 treatment wards due to an outbreak of the infection in the surrounding areas, interrupting the usual medical care for rehabilitation.^22,23)^ In contrast, the impact on inpatient rehabilitation in Hokkaido, Japan, in the early stages of the pandemic as shown in this study was limited. We were able to achieve good patient outcomes even with the reduced hours of rehabilitation training that came with COVID-19 restrictions. However, these outcomes achieved with the staffing constraints resulted in longer hours and physical and mental burnout for medical personnel. Our observations correspond closely with the reports from other epidemic areas of the world and need to be addressed in a world with COVID-19.^24)^

This study has some important limitations. First, the prevalence and impact of COVID-19 were dependent on regionality and national insurance systems. Second, this study was a retrospective observational design and was conducted at a single facility in the early epidemic area of Japan. Third, this emerging infectious disease is expected to last long. The results of this study reflect the situation during the very early stages of the pandemic, when PPEs were extremely scarce, and are descriptive of the response process and its results. The response to the upcoming waves of COVID-19 might improve significantly as the issues of low PPE supply and COVID-19 management strategy have improved over time.

In this study, we demonstrated that early COVID-19 pandemic in Hokkaido, Japan, did not negatively affect the qualitative outcomes of inpatient rehabilitation. This study suggests that it is possible to provide conventional rehabilitation outcomes with minimal negative influences during the pandemic if appropriate infection control measures are implemented.

## Data Availability

The data that support the findings of this study are available from the corresponding author, [author initials], upon reasonable request.

## Acknowledgments

We gratefully acknowledge all members of the infection control team in Aizen hospital for their dedicated work under the pandemic situation of COVID-19. The authors also thank Dr. Shuji Dohi, head of the hospital, for his kind support and advice.

## References

1. David S. Hui, Esam I Azhar, Tariq A. Madani, Francine Ntoumi, Richard Kock, Osman Dar, Giuseppe Ippolito, Timothy D Mchugh, Ziad A Memish, Christian Drosten, Alimuddin Zumla, Eskild Petersenet. The continuing 2019-nCoV epidemic threat of novel coronaviruses to global health—The latest 2019 novel coronavirus outbreak in Wuhan, China. Int J Infect Dis 2020;91:264–266. doi:10.1016/j.ijid.2020.01.009

2. World Health Organization: WHO Director-General’s opening remarks at the media briefing on COVID-19. 11 March, 2020. https://www.who.int/dg/speeches/detail/who-director-general-s-opening-remarks-at-the-media-briefing-on-covid-19---11-march-2020. Accessed 12 March 2020.

3. Yuki Furuse, Yura K Ko, Mayuko Saito, Yugo Shobugawa, Kazuaki Jindai, Tomoya Saito, Hiroshi Nishiura, Tomimasa Sunagawa, Motoi Suzuki, National Task Force for COVID-19 Outbreak in Japan. Epidemiology of COVID-19 Outbreak in Japan, January–March 2020. Jpn J Infect Dis 2020. doi:10.7883/yoken.JJID.2020.271

4. Shunji Edagawa, Fumiko Kobayashi, Fumihiro Kodama, Masayuki Takada, Yuki Itagaki, Akira Kodate, Keisuke Bando, Keisuke Sakurai, Akio Endo, Hisako Sageshima, Atsushi Nagasaka. Epidemiological features after emergency declaration in Hokkaido and report of 15 cases of COVID-19 including 3 cases requiring mechanical ventilation. Global Health & Medicine 2020; 2, 112–117. doi:10.35772/ghm.2020.01024

5. Mun-Keat Looi. Covid-19: Japan ends state of emergency but warns of “new normal”. BMJ 2020; 369: m2100. doi:10.1136/bmj.m2100

6. J M Linacre, AW Heinemann, B D Wright, C V Granger, B B Hamilton. The structure and stability of the Functional Independence Measure. Arch Phys Med Rehabil 1994 Feb;75(2):127–32.

7. Kanda Y. Investigation of the freely-available easy-to-use software “EZR” (Easy R) for medical statistics. Bone Marrow Transplant 2013:48, 452–458. doi:10.1038/bmt.2012.244

8. Information for Public. Q&A on Coronavirus Disease 2019 (COVID-19). Ministry of Health, Labor and Welfare.2020. https://www.mhlw.go.jp/stf/seisakunitsuite/bunya/kenkou_iryou/dengue_fever_qa_00014.html. Accessed 12 March 2020.

9. Ribeiro DKMN, Lenardt MH, Lourenço TM, Betiolli SE, Seima MD, Guimarães CA. The use of the functional independence measure in elderly. Rev Gaúcha Enferm 2017; 38(4): e66496. doi:10.1590/1983-1447.2017.04.66496

10. M. Tokunaga, S. Mita, Keiichi Tashiro, M. Yamaga, Y. Hashimoto, R. Nakanishi, H. Yamanaga. Methods for comparing Functional Independence Measure improvement degree for stroke patients between rehabilitation hospitals. Int J Phys Med Rehabil 2017;5:2. doi:10.4172/2329-9096.1000394

11. Centers for Disease Control and Prevention. Operational Considerations for the Identification of Healthcare Workers and Inpatients with Suspected COVID-19 in non-US Healthcare Settings. https://www.cdc.gov/coronavirus/2019-ncov/hcp/non-us-settings/guidance-identify-hcw-patients.html. Accessed: May 26 2020.

12. Turolla A, Rossettini G, Viceconti A, Palese A, Geri T. Musculoskeletal Physical Therapy During the COVID-19 Pandemic: Is Telerehabilitation the Answer? Phys Ther 2020 Aug 12;100(8):1260–1264. doi: 10.1093/ptj/pzaa093

13. Centers for Disease Control and Prevention. Infection Control Guidance for Healthcare Professionals about Coronavirus (COVID-19). https://www.cdc.gov/coronavirus/2019-ncov/hcp/infection-control.html. Accessed 11 March 2020.

14. Ferretti L, Wymant C, Kendall M, Zhao L, Nurtay A, Abeler-Dörner L, Parker M, Bonsall D, Fraser C. Quantifying SARS-CoV-2 transmission suggests epidemic control with digital contact tracing. Science 2020;368(6491):eabb6936. doi:10.1126/science.abb6936

15. Centers for Disease Control and Prevention. Protecting Healthcare Personnel. https://www.cdc.gov/hai/prevent/ppe.html. Accessed 26 May 2020.

16. Xiaoquan Lai, Minghuan Wang, Chuan Qin, Li Tan, Lusen Ran, Daiqi Chen, Han Zhang, Ke Shang, Chen Xia, Shaokang Wang, Shabei Xu, Wei Wang. Coronavirus Disease 2019 (COVID-2019) Infection Among Health Care Workers and Implications for Prevention Measures in a Tertiary Hospital in Wuhan, China. JAMA Netw Open. 2020 May 1;3(5):e209666. doi:10.1001/jamanetworkopen.2020.9666

17. Christopher M Petrilli, Simon A Jones, Jie Yang, Harish Rajagopalan, Luke O’Donnell, Yelena Chernyak, Katie A Tobin, Robert J Cerfolio, Fritz Francois, Leora I Horwitz. Factors associated with hospital admission and critical illness among 5279 people with coronavirus disease 2019 in New York City: prospective cohort study. BMJ 2020;369:m1966. doi:10.1136/bmj.m1966

18. Temet M McMichael, Dustin W Currie, Shauna Clark, Sargis Pogosjans, Meagan Kay, Noah G. Schwartz, James Lewis, Atar Baer, Vance Kawakami, Margaret D. Lukoff, Jessica Ferro, Claire Brostrom-Smith, Thomas D. Rea, Michael R. Sayre, Francis X. Riedo, Denny Russell, Brian Hiatt, Patricia Montgomery, Agam K. Rao, Eric J. Chow, Farrell Tobolowsky, Michael J. Hughes, Ana C. Bardossy, Lisa P. Oakley, Jesica R. Jacobs, Nimalie D. Stone, Sujan C. Reddy, John A. Jernigan, Margaret A. Honein, Thomas A. Clark, Jeffrey S. Duchin, for the Public Health– Seattle and King County, EvergreenHealth, and CDC COVID-19 Investigation Team. Epidemiology of Covid-19 in a Long-Term Care Facility in King County, Washington. N Engl J Med 2020 May 21;382(21):2005–2011. doi:10.1056/nejmoa2005412

19. Klompas M, Morris CA, Sinclair J, Pearson M, Shenoy ES. Universal masking in hospitals in the COVID-19 Era. N Engl J Med 2020;382:e63. doi:10.1056/nejmp2006372

20. Lindsley WG, Noti JD, Blachere FM, Szalajda JV, Beezhold DH. Efficacy of face shields against cough aerosol droplets from a cough simulator. J Occup Environ Hyg 2014;11(8):509–518. doi:10.1080/15459624.2013.877591

21. Perencevich EN, Diekema DJ, Edmond MB. Moving personal protective equipment into the community: face shields and containment of COVID-19. JAMA 2020;323(22):2252–2253. doi:10.1001/jama.2020.7477

22. Escalon MX, Herrera J. Adapting to the Coronavirus Disease 2019 Pandemic in New York City. Am J Phys Med Rehabil. 2020 Jun;99(6):453–458. doi:10.1093/ageing/afaa118

23. Paolo Boldrini, Carlotte Kiekens, Stefano Bargellesi, Rodolfo Brianti, Silvia Galeri, Lucia Lucca. First impact on services and their preparation. “Instant paper from the field” on rehabilitation answers to the Covid-19 emergency. Eur J Phys Rehabil Med 2020 Jun;56(3):319-322. doi.org/10.23736/s1973-9087.20.06303-0

24. Qian Liu, Dan Luo, Joan E Haase, Qiaohong Guo, Xiao Qin Wang, Shuo Liu, Lin Xia, Zhongchun Liu, Jiong Yang, Bing Xiang Yang. The experiences of health-care providers during the COVID-19 crisis in China: a qualitative study. Lancet Glob Health 2020 Jun;8(6):e790–e798. doi: 10.1016/S2214-109X(20)30204-7.

